# Heterogeneity of Treatment Effects of Glucose-lowering Drug Classes for Type 2 Diabetes: LEGEND-T2DM Network Real-World Evidence

**DOI:** 10.1101/2025.06.02.25328066

**Authors:** David M. Dávila-García, Thomas Falconer, Nicole Pratt, Karthik Natarajan, George Hripcsak

## Abstract

**Aims:** To assess heterogeneity of treatment effects (HTE) of glucose-lowering drug classes by clinical (cardiovascular [CV] risk, renal impairment) and demographic (age, sex) subgroups in adults with type 2 diabetes mellitus (T2D).

**Methods:** The LEGEND-T2DM network identified 4,746,939 adults with T2D on metformin monotherapy who initiated one of four glucose-lowering drug classes: glucagon-like peptide-1 receptor agonists (GLP-1 RA), sodium-glucose cotransporter 2 inhibitors (SGLT2i), dipeptidyl peptidase-4 inhibitor (DPP-4i) or sulfonylureas. HTE was assessed between glucose-lowering drug classes by clinical (low CV risk vs. higher CV risk; without renal impairment vs. renal impairment) and demographic (lower vs. middle vs. older age; male vs. female) subgroups. Outcomes included MACE (primary), acute myocardial infarction, stroke, sudden cardiac death and safety endpoints.

**Results:** Pairwise differences (n=1,115 tests) between adjusted hazard ratios (HRs) showed 49 nominally significant associations (p<0.05) and one statistically significant difference after Bonferroni correction (p<4.5×10^-5^). Among older subjects (vs. younger), those taking GLP-1 RA (vs. sulfonylureas) had statistically significant difference in risk of hypoglycemia (HRs: lower age, 0.53±0.14 vs. older age, 0.20±0.05, p<0.001).

**Conclusions:** HTE among glucose-lowering drug classes by clinical and demographic subgroups may provide guidance to generate hypothesis-testing studies to inform T2D treatment decisions.

## 1. INTRODUCTION

The prevalence of type 2 diabetes mellitus (T2D) continues to increase to epidemic proportions and remains a major cause of morbidity and mortality worldwide [1,2]. T2D is also the major cause of chronic kidney disease (CKD) and end-stage kidney disease [3]. While the presence of both T2D and CKD is associated with high mortality, cardiovascular disease (CVD) remains the primary cause of morbidity and mortality in individuals with T2D [4,5]. In the United States, the age-adjusted prevalence of total diabetes increased from 9.7% in 1999-2000 to 14.3% in 2021-2023, and glycemic control significantly declined after a decade of stability, raising concerns about an impending health crisis [6,7].

Recent randomized controlled trials (RCTs) have shown that new glucose-lowering drug classes (i.e., glucagon-like peptide-1 receptor agonist [GLP-1 RA], sodium-glucose cotransporter 2 inhibitor [SGLT2i]) improve glycemic control, reduce risk of major adverse cardiovascular events (MACE) and decrease progression of CKD [8,9]. Dipeptidyl peptidase-4 inhibitors (DPP-4is) also improve glycemic control, but have not been shown to lower risk of MACE in several RCTs [10–13]. Sulfonylureas, despite being an older class of glucose-lowering agents, remain widely used due to their efficacy and affordability. However, this drug class is associated with increased risk of hypoglycemia and weight gain [14]. Together, these glucose-lowering drug classes have become standard of care in the guidelines of major associations including American Diabetes Association and the European Association for the Study of Diabetes [9,15].

A recent study from the Large-scale Evidence Generation and Evaluation across a Network of Databases for Type 2 Diabetes Mellitus (LEGEND-T2DM) network assessed comparative effectiveness of these classes in head-to-head analyses, using data from over 1.5 million patients with T2D and established CVD treated with metformin [16]. Their study evaluated 3-point MACE (acute myocardial infarction, stroke and sudden cardiac death), 4-point MACE (3-point MACE plus hospitalization for heart failure) and their individual components. They found that GLP-1 RA and SGLT2i reduced the risk for 3- and 4-point MACE compared with sulfonylureas and DPP-4i [16,17]. Furthermore, relative to sulfonylureas, DPP-4i reduced the risk of 3-point MACE (HR: 0.87; 95% CI: 0.79-0.95), which is in disagreement with RCTs [18,19]. No differences were observed between SGLT2i and GLP-1 RA for 3- or 4-point MACE, or hospitalization for heart failure. However, relative to GLP-1 RA, SGLT2i were associated with increased risk of acute myocardial infarction (HR: 1.19; 95% CI: 1.05-1.35).

Evaluations of heterogeneity of treatment effects and safety profiles with respect to glucose-lowering drug classes using real-world data remain limited. Bridging this knowledge gap may guide the development of hypothesis-driven research that informs personalized treatment decisions in T2D. A recent study on 23,137 Scottish patients with newly diagnosed T2D showed not only that underlying phenotypic variation drives T2D onset, but that it also affects subsequent outcomes and drug response, “demonstrating the need to incorporate these factors into personalized treatment approaches” for proper management [20]. Accordingly, we leveraged data from the LEGEND-T2DM study, which comprises a multinational network of 10 administrative claims and electronic health record databases containing over 4.7 million records of adults with T2D. The overarching aim of this study was to assess the heterogeneity of treatment effects and safety profiles among pairwise glucose-lowering drug classes across broad clinical (cardiovascular risk, renal impairment) and demographic (age, sex) subgroups.

## 2. MATERIALS AND METHODS

The LEGEND-T2DM study leveraged electronic health records and administrative claims collected through the Observational Health Data Sciences and Informatics network to perform systematic, large-scale, multinational real-world comparative cardiovascular effectiveness and safety studies of four major glucose-lowering drug classes (SGLT2i, GLP-1 RA, DPP-4i and sulfonylureas) [16,21]. The data were obtained for analysis from the publicly available LEGEND-T2DM study R Shiny application on September 26, 2024 [22]. Detailed study methods have been previously reported and modeled after the LEGEND-HTN study [16,21,23–25]. The present study was approved by the Columbia University Institutional Review Board (approval date: December 2024; reference number AAAO7805); patient informed consent was waived.

### 2.1 Database Information

The LEGEND-T2DM network included 10 federated longitudinal data sources across 4 countries (United States, Germany, Spain, and the United Kingdom), yielding over 4.7 million subjects who met inclusion criteria. The data sources included 6 administrative claims databases and 4 electronic health records databases. US populations included those commercially and publicly insured, enriched for older individuals (i.e., Medicare and Veterans Affairs), lower socioeconomic status (i.e., Medicaid), and racially diverse (e.g., Veterans Affairs >20% Black or African American) [24]. All data sources were mapped to the Observational Medical Outcomes Partnership (OMOP) common data model to improve standardization and interoperability [26]. Individual data source characteristics for this analysis were previously reported [16]. Further details regarding concept set definitions for exposures, outcomes, subgroups and population cohorts were previously described by Khera et al. [16,21].

### 2.2 Exposures

Pairwise (n=6) class-level exposure comparisons of the four major glucose-lowering drug classes (i.e., GLP-1 RA, SGLT2i, DPP-4i, sulfonylureas) were performed to estimate hazard ratios across all 32 study outcomes. Continuous drug exposure was defined as consecutive drug prescriptions within a 30-day window. An on-treatment (also referred to as “per-protocol”) time-at-risk definition was used: subjects were followed from index date to treatment discontinuation [27]. The on-treatment analysis accounts for direct treatment effects while accommodating treatment escalation with additional glucose-lowering drugs [28]. Thiazolidinediones were excluded due to the known association with risk of heart failure and bladder cancer [29].

### 2.3 Study Population

The LEGEND-T2DM study population comprised adults (≥18 years) diagnosed with T2D with a history of prior metformin monotherapy, who were initiated on treatment with any one of 22 major glucose-lowering drug agents from the DPP-4i, GLP-1 RA, SGLT2i or sulfonylureas classes, as previously reported by Khera et al. [21]. Study inclusion criteria: a) T2D diagnosis; b) ≥1 year of observation prior to index date; c) ≥3 months of metformin use prior to index date. Exclusion criteria: a) prior diagnoses of type 1 or secondary diabetes; b) prior exposures to DPP-4i, GLP-1 RA, SGLT2i, sulfonylureas or any other glucose-lowering drug agents or >30 days of insulin exposure prior to the index date. Patients with higher CV risk or renal impairment were included to characterize the heterogeneity of treatment effects and safety profiles of these agents across these known comorbidities. The index date was determined for each patient as the first observed exposure to any one of the four glucose-lowering drug classes.

### 2.4 Clinical and demographic subgroups

The study population was stratified into univariate clinical and demographic subgroups to assess heterogeneity of treatment effects. Across all data sources, HRs for all 32 outcomes were estimated for all pairwise class-level exposure cohorts and all subgroups. We identified patients with higher CV risk by adopting an established CVD definition previously validated among new-users of glucose-lowering drugs [30]. Established CVD was defined as either having at least one diagnosis code for a condition indicating CVD (e.g. atherosclerotic vascular disease, cerebrovascular disease, ischemic heart disease, or peripheral vascular disease) or having undergone at least one procedure indicating CVD (e.g. percutaneous coronary intervention, coronary artery bypass graft or revascularization). Similarly, renal impairment was defined using diagnosis codes for chronic kidney disease and end-stage renal disease, dialysis procedures or abnormal laboratory measures indicative of renal dysfunction (e.g., serum creatinine, estimated glomerular filtration rate or urine albumin).

Heterogeneity of treatments were assessed for primary cardiovascular, secondary effectiveness, and secondary patient-centered safety outcomes across subgroups. Our approach aimed to determine whether significant differences in hazard ratio estimates exist between drug classes across clinical and demographic subgroups. Clinical subgroups included: “cardiovascular risk”—low cardiovascular risk (LCVR) vs. higher cardiovascular risk (HCVR); and “renal impairment”—without renal impairment (WRI) vs. renal impairment (RI). Demographic subgroups included: “age groups”—i. lower age (18–44 years) vs. middle age (45–64 years); ii. lower age vs. older age (≥65 years); iii. middle age vs. older age; and “biological sex”—male vs. female.

### 2.5 Study outcomes

The primary outcomes included the composite CV endpoints of 3- and 4-point MACE. Secondary outcomes encompassed individual CV effectiveness endpoints, while secondary patient-centered outcomes addressed safety concerns. Subjects who experienced an outcome prior to their index date were excluded from the exposure cohorts used to generate HR estimates.

Calibrated HRs used in this analysis were derived from previously reported LEGEND-T2DM results, which performed propensity score matching and random-effects meta-analysis with empirical calibration. Additional methodological details are available in the original LEGEND-T2DM publication and summarized in the Supplemental Methods [16,21].

### 2.6 Statistical analysis

Statistical approaches to evaluate heterogeneity of treatment effects and safety profiles across the clinical (cardiovascular risk, renal impairment) and demographic (age, sex) subgroups were used. Across all data sources and pairwise exposure cohorts, the meta-analysis-calibrated HRs and corresponding standard errors (SEs) for all 32 outcomes were estimated [16,21]. The hazard, ℎ(𝑡), is defined as the instantaneous rate of experiencing an outcome at time 𝑡, conditional on having not experienced the outcome prior to time 𝑡. The HR represents the relative hazard of experiencing an outcome at time 𝑡 between individuals receiving the target vs. comparator drug classes (Equation 1) [31]. We assumed that log-transformed HR estimates follow an asymptotically normal distribution [32].

To quantify heterogeneity of treatment effects across subgroups, we calculated the difference between log-transformed hazard ratios, denoted as Δln(𝐻*R*), obtained between subgroups within the same drug-outcome comparison (Equation 2). The corresponding standard error associated with this difference (𝑆*E*_Δln(𝐻*R*)_) was computed using the subgroup-specific SEs (Equation 3). The statistical significance of subgroup differences was determined by calculating a Z-score test statistic, defined as the ratio of the difference in log-transformed HRs to its standard error (Equation 4).

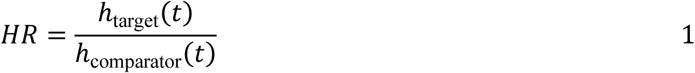

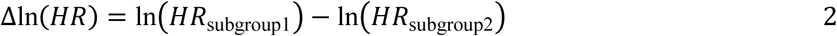

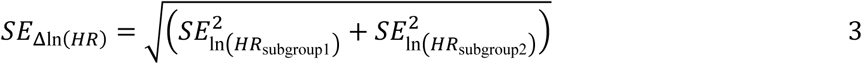

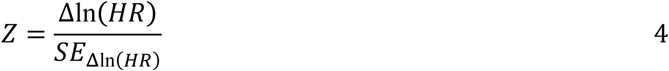

Subgroup differences in drug class comparisons were considered nominally significant if the p-value was less than 0.05. The analysis involved 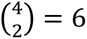 unique drug class exposure pairs, 6 subgroup comparisons and 32 clinical outcomes. This design allowed for a maximum of 6 × 6 × 32 = 1,152 hypothesis tests, of which 37 (3.2%) could not be performed due to missing data, yielding 1,115 completed tests. To adjust for multiple hypothesis testing, a Bonferroni correction was applied 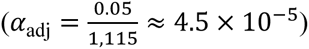; thus, only differences with p-values less than 𝛼_adj_ were considered statistically significant.

## 3. RESULTS

### 3.1 Cohort Characteristics

There was a total of 4,746,939 subjects with T2D across the 10 federated data sources from 4 countries. Among these individuals, SGLT2is were initiated in 1,186,078 (25.0%), GLP-1 RAs in 384,201 (8.1%), DPP-4is in 738,880 (15.5%), and sulfonylureas in 2,441,944 (51.4%). Overall, the subgroup proportions by drug classes were as follows: HCVR, 26.9–32.6%; RI, 8.6–12.4%; lower age, 10.8–12.8%; middle age, 48.3–59.1%; older age, 23.6–40.3%; female, 44.7–56.0% (Table 1). Calibrated HR estimates with nominally or statistically significant differences are shown by group: cardiovascular risk subgroups (Table 2), renal impairment subgroups (Table 3), age groups (Table 4A-C), and sex (Table 5). Differences in the primary endpoint (i.e., MACE) and secondary effectiveness outcomes (i.e., acute myocardial infarction, acute renal failure, glycemic control, hospitalization for heart failure, measured renal dysfunction, revascularization, stroke and sudden cardiac death) are reported. Significant differences in safety outcomes are provided in Tables 2-5. Out of 1,115 pairwise tests conducted, 49 (4.4%) nominally significant differences in treatment effectiveness or safety outcomes across T2D clinical and demographic subgroups were found, of which only 1 remained statistically significant after Bonferroni correction. The tables include additional findings that may inform future hypothesis-generating explorations.

**Table 1.** Cohort size by drug class and subgroups (n=4,746,939)

| Table 1. Cohort size by drug class and subgroups (n=4,746,939) |  |  |  |  |
| --- | --- | --- | --- | --- |
| Subgroup | Glucose-lowering drug classes |  |  |  |
|  | SGLT2i<br>(n=1,186,078) | GLP-1 RA<br>(n=384,201) | DPP-4i<br>(n=734,716) | Sulfonylureas<br>(n=2,441,944) |
| **Cardiovascular risk subgroups** |  |  |  |  |
| Low CVR | 799,770 (67.4%) | 280,994 (73.1%) | 492,130 (69.2%) | 1,653,538 (67.7%) |
| Higher CVR | 386,308 (32.6%) | 103,207 (26.9%) | 218,987 (30.8%) | 788,406 (32.3%) |
| Totals | 1,186,078 | 384,201 | 711,117 | 2,441,944 |
| **Kidney function subgroups** |  |  |  |  |
| Without renal impairment | 1,034,145 (88.8%) | 351,138 (91.4%) | 669,571 (91.1%) | 2,015,852 (87.6%) |
| Renal impairment | 130,467 (11.2%) | 32,955 (8.6%) | 65,145 (8.9%) | 286,446 (12.4%) |
| Totals | 1,164,612 | 384,093 | 734,716 | 2,302,298 |
| **Age subgroups** |  |  |  |  |
| Lower age (18-44y) | 126,507 (10.8%) | 65,933 (17.3%) | 91,236 (12.8%) | 268,520 (11.4%) |
| Middle age (45-64y) | 593,322 (50.6%) | 225,135 (59.1%) | 420,976 (59.1%) | 1,137,871 (48.3%) |
| Older age (≥65y) | 452,409 (38.6%) | 89,853 (23.6%) | 200,254 (28.1%) | 949,021 (40.3%) |
| Totals | 1,172,238 | 380,921 | 712,466 | 2,355,412 |
| **Sex subgroups** |  |  |  |  |
| Male | 581,812 (50.1%) | 167,744 (44.0%) | 396,532 (55.3%) | 1,187,739 (52.1%) |
| Female | 579,996 (49.9%) | 213,533 (56.0%) | 320,709 (44.7%) | 1,093,069 (47.9%) |
| Totals | 1,161,808 | 381,277 | 717,241 | 2,280,808 |
| CVR: cardiovascular risk, DPP-4i: Dipeptidyl peptidase-4 inhibitors, GLP-1 RA: Glucagon-like peptide-1 receptor agonist, SGLT2i: Sodium-glucose co-transporter-2 inhibitors. |  |  |  |  |

**Table 2.** Significant HR Differences: Low CVR vs. Higher CVR Subgroups.

| Table 2. Significant HR Differences: Low CVR vs. Higher CVR Subgroups |  |  |  |  |  |
| --- | --- | --- | --- | --- | --- |
| Outcome | Target Drug Class | Comparator Drug Class | HR 1 (low CVR) | HR 2 (higher CVR) | p-value |
| 3-point MACE | DPP-4i | SGLT2i | **1.05 ± 0.11** | **1.23 ± 0.12** | **0.027** |
| Sudden Cardiac Death | DPP-4i | SGLT2i | **1.17 ± 0.36** | **1.69 ± 0.46** | **0.047** |
| Glycemic Control | DPP-4i | SGLT2i | **0.97 ± 0.12** | **1.19 ± 0.21** | **0.036** |
| Hyperkalemia | DPP-4i | SGLT2i | 1.01 ± 0.22 | 1.42 ± 0.28 | 0.013 |
| Peripheral Edema | DPP-4i | GLP-1 RA | 1.07 ± 0.12 | 1.25 ± 0.14 | 0.042 |
| Bladder Cancer | GLP-1 RA | SGLT2i | 1.74 ± 0.97 | 0.88 ± 0.50 | 0.035 |
| Hazard ratio estimates (HRs) from individual data sources were aggregated using random effects meta-analysis, followed by calibration using empirical null distributions to correct for residual confounding and systematic bias. Only bolded pairwise differences are discussed in the text.<br>CVR: cardiovascular risk, DPP-4i: dipeptidyl peptidase-4 inhibitor(s), GLP-1 RA: glucagon-like peptide-1 receptor agonist(s), HR: hazard ratio, MACE: major adverse cardiovascular events, SGLT2i: sodium-glucose cotransporter 2 inhibitor(s), SU: sulfonylureas |  |  |  |  |  |

**Table 3.** Significant HR Differences: Without Renal Impairment vs. Renal Impairment Subgroups.

|  |  |  |  |  |  |
| --- | --- | --- | --- | --- | --- |
| **Outcome** | **Target Drug Class** | **Comparator Drug Class** | **HR 1 (Without RI)** | **HR 2 (RI)** | **p-value** |
| **Sudden Cardiac Death** | **DPP-4i** | **SU** | **0.84 ± 0.12** | **1.05 ± 0.17** | **0.029** |
| **Stroke** | **DPP-4i** | **SGLT2i** | **1.03 ± 0.22** | **1.41 ± 0.36** | **0.040** |
| All-cause Mortality | DPP-4i | SGLT2i | 1.55 ± 0.50 | 0.74 ± 0.56 | 0.022 |
| Hyperkalemia | DPP-4i | SGLT2i | 1.18 ± 0.15 | 1.68 ± 0.34 | 0.002 |
| Abnormal Weight Gain | DPP-4i | SGLT2i | 1.09 ± 0.17 | 1.96 ± 0.92 | 0.006 |
| Vomiting | DPP-4i | SGLT2i | 1.02 ± 0.11 | 1.26 ± 0.25 | 0.045 |
| Thyroid Tumor | GLP-1 RA | SGLT2i | 0.68 ± 0.30 | 1.68 ± 1.78 | 0.029 |
| Peripheral Edema | DPP-4i | GLP-1 RA | 1.15 ± 0.12 | 1.39 ± 0.22 | 0.038 |
| Peripheral Edema | GLP-1 RA | SU | 0.82 ± 0.17 | 0.63 ± 0.08 | 0.022 |
| Hypoglycemia | GLP-1 RA | SU | 0.37 ± 0.09 | 0.20 ± 0.06 | <0.001 |
| Hazard ratio estimates (HRs) from individual data sources were aggregated using random effects meta-analysis, followed by calibration using empirical null distributions to correct for residual confounding and systematic bias. Only bolded pairwise differences are discussed in the text.<br>DPP-4i: dipeptidyl peptidase-4 inhibitor(s), GLP-1 RA: glucagon-like peptide-1 receptor agonist(s), HR: hazard ratio, RI: renal impairment, SGLT2i: sodium-glucose cotransporter 2 inhibitor(s), SU: sulfonylureas |  |  |  |  |  |

**Table 4.**
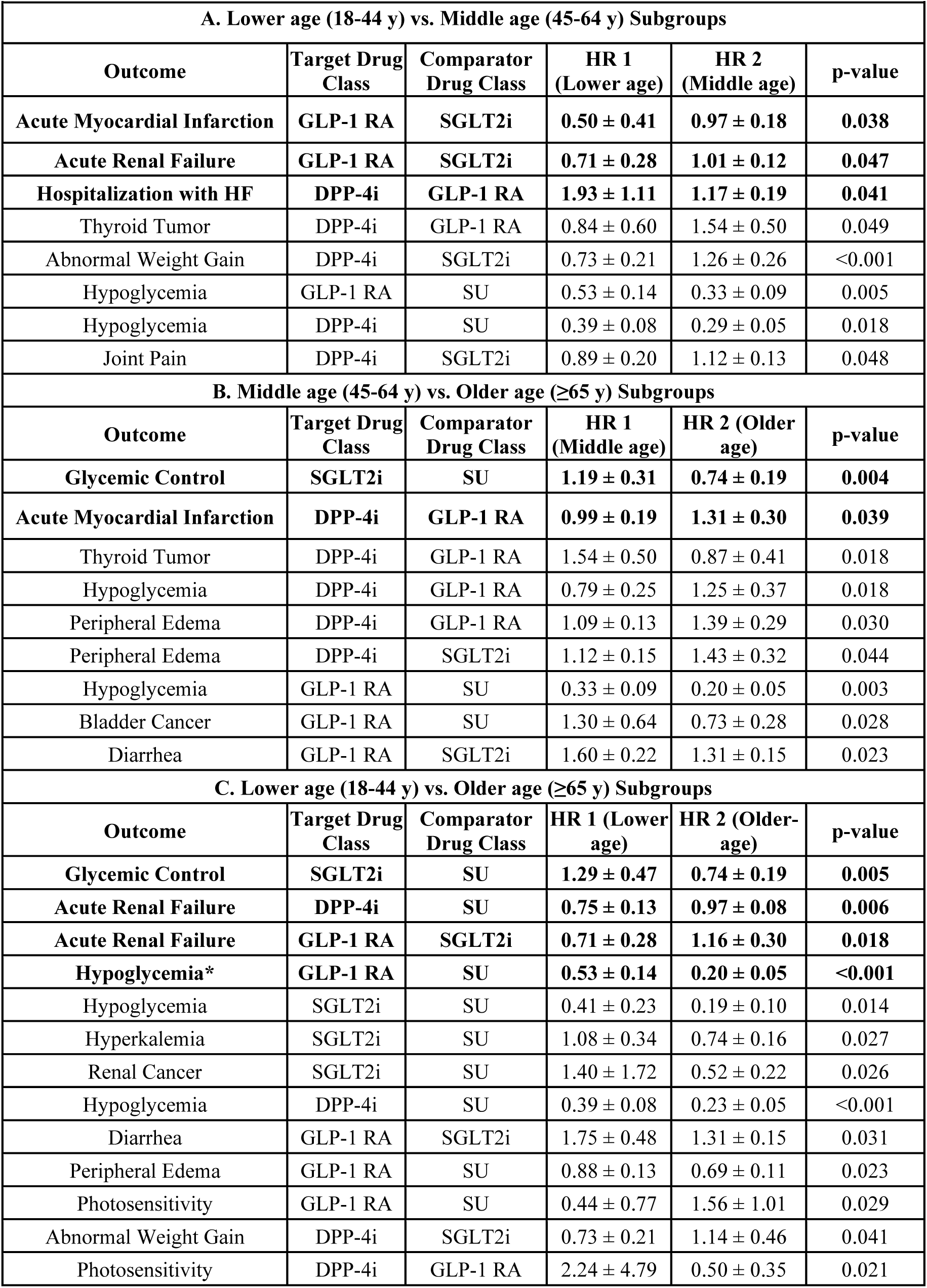
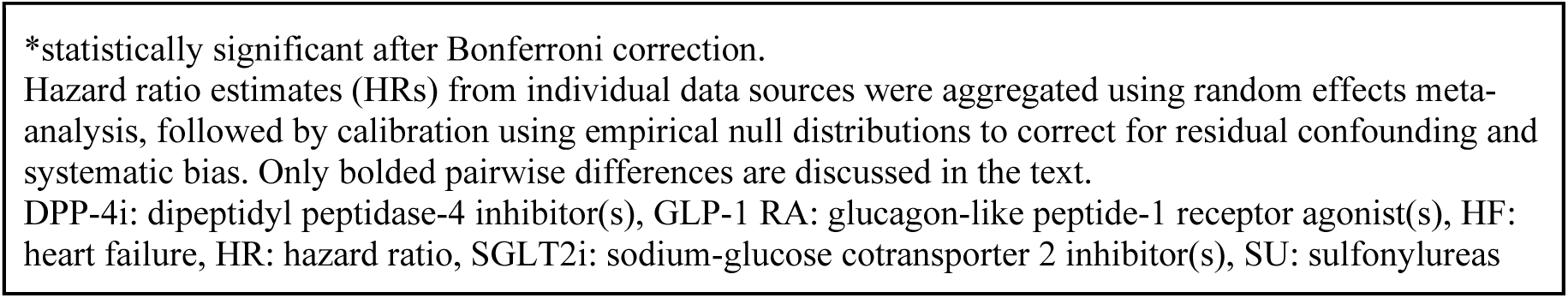
Significant HR Differences: Age Subgroups.

| **Table 4. Significant HR Differences: Age Subgroups** |  |  |  |  |  |
| --- | --- | --- | --- | --- | --- |
| **A. Lower age (18-44 y) vs. Middle age (45-64 y) Subgroups** |  |  |  |  |  |
| **Outcome** | **Target Drug Class** | **Comparator Drug Class** | **HR 1 (Lower age)** | **HR 2 (Middle age)** | **p-value** |
| **Acute Myocardial Infarction** | **GLP-1 RA** | **SGLT2i** | **0.50 ± 0.41** | **0.97 ± 0.18** | **0.038** |
| **Acute Renal Failure** | **GLP-1 RA** | **SGLT2i** | **0.71 ± 0.28** | **1.01 ± 0.12** | **0.047** |
| **Hospitalization with HF** | **DPP-4i** | **GLP-1 RA** | **1.93 ± 1.11** | **1.17 ± 0.19** | **0.041** |
| Thyroid Tumor | DPP-4i | GLP-1 RA | 0.84 ± 0.60 | 1.54 ± 0.50 | 0.049 |
| Abnormal Weight Gain | DPP-4i | SGLT2i | 0.73 ± 0.21 | 1.26 ± 0.26 | <0.001 |
| Hypoglycemia | GLP-1 RA | SU | 0.53 ± 0.14 | 0.33 ± 0.09 | 0.005 |
| Hypoglycemia | DPP-4i | SU | 0.39 ± 0.08 | 0.29 ± 0.05 | 0.018 |
| Joint Pain | DPP-4i | SGLT2i | 0.89 ± 0.20 | 1.12 ± 0.13 | 0.048 |
| **B. Middle age (45-64 y) vs. Older age (≥65 y) Subgroups** |  |  |  |  |  |
| **Outcome** | **Target Drug Class** | **Comparator Drug Class** | **HR 1 (Middle age)** | **HR 2 (Older age)** | **p-value** |
| **Glycemic Control** | **SGLT2i** | **SU** | **1.19 ± 0.31** | **0.74 ± 0.19** | **0.004** |
| **Acute Myocardial Infarction** | **DPP-4i** | **GLP-1 RA** | **0.99 ± 0.19** | **1.31 ± 0.30** | **0.039** |
| Thyroid Tumor | DPP-4i | GLP-1 RA | 1.54 ± 0.50 | 0.87 ± 0.41 | 0.018 |
| Hypoglycemia | DPP-4i | GLP-1 RA | 0.79 ± 0.25 | 1.25 ± 0.37 | 0.018 |
| Peripheral Edema | DPP-4i | GLP-1 RA | 1.09 ± 0.13 | 1.39 ± 0.29 | 0.030 |
| Peripheral Edema | DPP-4i | SGLT2i | 1.12 ± 0.15 | 1.43 ± 0.32 | 0.044 |
| Hypoglycemia | GLP-1 RA | SU | 0.33 ± 0.09 | 0.20 ± 0.05 | 0.003 |
| Bladder Cancer | GLP-1 RA | SU | 1.30 ± 0.64 | 0.73 ± 0.28 | 0.028 |
| Diarrhea | GLP-1 RA | SGLT2i | 1.60 ± 0.22 | 1.31 ± 0.15 | 0.023 |
| **C. Lower age (18-44 y) vs. Older age (≥65 y) Subgroups** |  |  |  |  |  |
| **Outcome** | **Target Drug Class** | **Comparator Drug Class** | **HR 1 (Lower age)** | **HR 2 (Older age)** | **p-value** |
| **Glycemic Control** | **SGLT2i** | **SU** | **1.29 ± 0.47** | **0.74 ± 0.19** | **0.005** |
| **Acute Renal Failure** | **DPP-4i** | **SU** | **0.75 ± 0.13** | **0.97 ± 0.08** | **0.006** |
| **Acute Renal Failure** | **GLP-1 RA** | **SGLT2i** | **0.71 ± 0.28** | **1.16 ± 0.30** | **0.018** |
| **Hypoglycemia\*** | **GLP-1 RA** | **SU** | **0.53 ± 0.14** | **0.20 ± 0.05** | **<0.001** |
| Hypoglycemia | SGLT2i | SU | 0.41 ± 0.23 | 0.19 ± 0.10 | 0.014 |
| Hyperkalemia | SGLT2i | SU | 1.08 ± 0.34 | 0.74 ± 0.16 | 0.027 |
| Renal Cancer | SGLT2i | SU | 1.40 ± 1.72 | 0.52 ± 0.22 | 0.026 |
| Hypoglycemia | DPP-4i | SU | 0.39 ± 0.08 | 0.23 ± 0.05 | <0.001 |
| Diarrhea | GLP-1 RA | SGLT2i | 1.75 ± 0.48 | 1.31 ± 0.15 | 0.031 |
| Peripheral Edema | GLP-1 RA | SU | 0.88 ± 0.13 | 0.69 ± 0.11 | 0.023 |
| Photosensitivity | GLP-1 RA | SU | 0.44 ± 0.77 | 1.56 ± 1.01 | 0.029 |
| Abnormal Weight Gain | DPP-4i | SGLT2i | 0.73 ± 0.21 | 1.14 ± 0.46 | 0.041 |
| Photosensitivity | DPP-4i | GLP-1 RA | 2.24 ± 4.79 | 0.50 ± 0.35 | 0.021 |

\*statistically significant after Bonferroni correction.
Hazard ratio estimates (HRs) from individual data sources were aggregated using random effects meta-analysis, followed by calibration using empirical null distributions to correct for residual confounding and systematic bias. Only bolded pairwise differences are discussed in the text.
DPP-4i: dipeptidyl peptidase-4 inhibitor(s), GLP-1 RA: glucagon-like peptide-1 receptor agonist(s), HF: heart failure, HR: hazard ratio, SGLT2i: sodium-glucose cotransporter 2 inhibitor(s), SU: sulfonylureas

**Table 5.** Significant HR Differences: Male vs. Female Subgroups.

| Table 5. Significant HR Differences: Male vs. Female Subgroups |  |  |  |  |  |
| --- | --- | --- | --- | --- | --- |
| Outcome | Target Drug Class | Comparator Drug Class | HR 1 (Male) | HR 2 (Female) | p-value |
| Glycemic Control | DPP-4i | SGLT2i | **0.88 ± 0.16** | **1.13 ± 0.17** | **0.023** |
| Abnormal Weight Loss | DPP-4i | SGLT2i | 0.80 ± 0.09 | 0.94 ± 0.10 | 0.025 |
| Acute Pancreatitis | GLP-1 RA | SU | 0.72 ± 0.20 | 1.04 ± 0.30 | 0.036 |
| Hazard ratio estimates (HRs) from individual data sources were aggregated using random effects meta-analysis, followed by calibration using empirical null distributions to correct for residual confounding and systematic bias. Only bolded pairwise differences are discussed in the text.<br>DPP-4i: dipeptidyl peptidase-4 inhibitor(s), GLP-1 RA: glucagon-like peptide-1 receptor agonist(s), HR: hazard ratio, SGLT2i: sodium-glucose cotransporter 2 inhibitor(s), SU: sulfonylureas |  |  |  |  |  |

### 3.2 Low vs. Higher Cardiovascular Risk Subgroups (Table 2)

Among subjects with HCVR (vs. LCVR), those taking DPP-4i (vs. SGLT2i) had increased difference in risk of “3-point MACE” (HRs: LCVR, 1.05±0.11 vs. HCVR, 1.23±0.12, p<0.027) and “sudden cardiac death” (HRs: LCVR, 1.17±0.36 vs. HCVR, 1.69±0.46, p<0.047). Additionally, for individuals with HCVR (vs. LCVR), those taking DPP-4i (vs. SGLT2i) exhibited improved “glycemic control” (HRs: LCVR, 0.97±0.12 vs. HCVR, 1.19±0.21, p<0.036).

### 3.3 Renal Impairment vs. Without Renal Impairment Subgroups (Table 3)

Among subjects with RI (vs. WRI), those taking DPP-4i (vs. sulfonylureas) had increased difference in risk of “sudden cardiac death” (HRs: WRI, 0.84±0.12 vs. RI, 1.05±0.17, p<0.029). For subjects with RI (vs. WRI), those who initiated DPP-4i (vs. SGLT2i) had increased difference in risk of “stroke” (HRs: WRI, 1.03±0.22 vs. RI, 1.41±0.36, p<0.040).

### 3.4 Lower vs. Middle vs. Older Age Subgroups (Table 4A-C)

#### 3.4.1 GLP-1 RA vs. SGLT2i

Among middle-aged subjects (vs. lower-aged), those taking GLP-1 RA (vs. SGLT2i) had increased difference in risk of “acute myocardial infarction” (HRs: lower age, 0.50±0.41 vs. middle age, 0.97 ± 0.18, p<0.038). Among older subjects (vs. younger), those taking GLP-1 RA (vs. SGLT2i) had higher risk of “acute renal failure”, which increased with age (HRs: lower age, 0.71±0.28 vs. middle age, 1.01±0.12, p<0.047; and lower age, 0.71±0.28 vs. older age, 1.16±0.30, p<0.018).

#### 3.4.2 DPP-4i vs. GLP-1 RA

Among middle-aged subjects (vs. lower-aged), those taking DPP-4i (vs. GLP-1 RA) had elevated difference in risk of “hospitalization with heart failure” (HRs: lower age, 1.93±1.11 vs. middle age, 1.17±0.19, p<0.041).

Among older-aged subjects (vs. middle-aged), those taking DPP-4i (vs. GLP-1 RA) had higher risk of “acute myocardial infarction” (HRs: middle age, 0.99±0.19 vs. older age, 1.31±0.30, p<0.039).

#### 3.4.3 GLP-1 RA vs. sulfonylureas

Among older subjects (vs. younger), those taking GLP-1 RA (vs. sulfonylureas) had statistically significant difference in risk of “hypoglycemia” (HRs: lower age, 0.53±0.14 vs. middle age, 0.33±0.09, p<0.005; middle age, 0.33±0.09 vs. older age, 0.20±0.05, p<0.003; and lower age, 0.53±0.14 vs. older age, 0.20±0.05, p<0.001). This was the only finding that remained statistically significant after Bonferroni correction.

#### 3.4.4 DPP-4i vs. sulfonylureas

Among older-aged subjects (vs. lower-aged), those taking DPP-4i (vs. sulfonylureas) had increased difference in risk of “acute renal failure” (HRs: lower age, 0.75±0.13 vs. older age, 0.97±0.08, p<0.006).

#### 3.4.5 SGLT2i vs. sulfonylureas

Among older subjects (vs. younger), those taking SGLT2i (vs. sulfonylureas) had decreased “glycemic control” (HRs: middle age, 1.19±0.31 vs. older age, 0.74±0.19, p<0.004; and lower age, 1.29±0.47 vs. older age, 0.74±0.19, p<0.005).

### 3.5 Male vs. Female Sex Subgroups (Table 5)

Among female subjects (vs. male), those taking DPP-4i (vs. SGLT2i) exhibited improved “glycemic control” (HRs: male, 0.88±0.16 vs. female, 1.13±0.17, p<0.023).

## 4. DISCUSSION

In this study, we conducted a systematic assessment of heterogeneity of treatment effects and safety profiles across four major glucose-lowering drug classes, using real-world data from over 4.7 million adults with T2D in the LEGEND-T2DM network. To our knowledge, this is the largest hypothesis-generating heterogeneity of treatment effect analysis of adults with T2D on metformin monotherapy who initiated one of four major glucose-lowering drug classes (GLP-1 RA, SGLT2i, DPP-4i or sulfonylureas). By analysis of adjusted hazard ratio differences, we outlined a statistical framework to assess heterogeneity in treatment effects across clinical and demographic subgroups. Unlike prior investigations that have largely focused on average treatment effects, our analysis examined how clinical (cardiovascular risk, renal impairment) and demographic (age, sex) patient subgroups modify the relative benefits and safety profiles of the glucose-lowering drug classes. Out of 1,115 pairwise tests conducted, 49 (4.4%) exhibited nominally significant differences, and only one exhibited a statistically significant difference after Bonferroni correction. Altogether, these findings suggest that comorbidity profiles at baseline may contribute to heterogeneous treatment responses. Given the distinct risk-benefit patterns observed across different subgroups, as further described below, our study confirms findings from previous studies. Furthermore, our findings suggest that hypothesis-testing studies based on the findings of the current study may further inform guidelines on the use of glucose-lowering drug classes.

### 4.1 Cardiovascular Risk Subgroup Heterogeneity

We found that those with higher (vs. low) CV risk taking DPP-4i (vs. SGLT2i) exhibited increased difference in risk of developing “3-point MACE” (acute myocardial infarction, stroke, sudden cardiac death) and “sudden cardiac death”. Those with low CV risk taking DPP-4i (vs. SGLT2i) exhibited no evidence of a difference in treatment effect for “3-point MACE” or “sudden cardiac death”. Recent RCTs show that SGLT2i significantly reduces the risk of MACE, heart failure and cardiovascular complications, regardless of baseline CV risk. Furthermore, DPP-4i are generally considered to have a neutral effect on cardiovascular outcomes [10,33,34].

For subjects with higher (vs. low) CV risk, those taking DPP-4i (vs. SGLT2i) had improved “glycemic control”. Those at low CV risk taking DPP-4i (vs. SGLT2i) exhibited non-inferior “glycemic control”. Despite the well-known importance of glycemic control to prevent CVD, we found no studies comparing these two drug classes by CV subgroups, representing a knowledge gap.

### 4.2 Renal Impairment Subgroup Heterogeneity

The latest clinical guidelines recommend initiation of metformin and SGLT2i as first-line treatments for glycemic management in patients with T2D and kidney disease [3]. Our findings support this recommendation. We found that subjects with renal impairment taking DPP-4i (vs. SGLT2i) exhibited higher risk of “stroke”; no evidence of a difference in treatment effect was observed for those without renal impairment. This finding aligns with current clinical guidelines recommending the use of SGLT2i over DPP-4i for patients with CKD, as SGLT-2i provide both cardiovascular and renal protection [3,35]. We also found that subjects with renal impairment taking DPP-4i (vs. sulfonylureas) had no evidence of a difference in treatment effect for “sudden cardiac death”. Furthermore, those without renal impairment on DPP-4i (vs. sulfonylureas) had lower risk for “sudden cardiac death”. DPP-4i have previously been shown to be preferred to sulfonylureas for low-risk patients (i.e., low CVR and without CKD) initiating glucose-lowering treatment [36].

### 4.3 Age Subgroup Heterogeneity

We found varying risk profiles for subjects treated with GLP-1 RA compared to SGLT2i, stratified by age group. For middle-aged subjects, those taking GLP-1 RA (vs. SGLT2i) showed no evidence of a difference in treatment effect for “acute myocardial infarction”, whereas lower-aged subjects taking GLP-1 RA (vs. SGLT2i) experienced decreased risk. Regarding “acute renal failure”, the hazard associated with GLP-1 RA (vs. SGLT2i) varied by age group: older-aged subjects had increased risk, middle-aged subjects showed no evidence of a difference in treatment effect, and lower-aged subjects had a decreased risk. For our study population, these findings suggest that while GLP-1 RAs were favored for younger adults with T2D, SGLT2i drugs were favored over GLP-1 RAs in older adults with T2D [37,38]. A recent systematic review and meta-analysis of 11 RCTs have shown that GLP-1 RAs reduce MACE, CV deaths, stroke and myocardial infarction to the same degree for people over and under 65 years of age [38,39]. Stratified analyses from the CANVAS Program, EMPA-REG OUTCOME and DECLARE–TIMI 58 RCTs suggest that older adults with T2D who initiated SGLT2i exhibited similar or greater benefits compared to younger adults with T2D, providing further evidence in support of our findings [37,40–42]. We observed significant risk differences by age group for subjects treated with DPP-4i compared to GLP-1 RA. For middle-aged subjects, those taking GLP-1 RA (vs. DPP-4i) showed no difference in treatment effect for “hospitalization for heart failure”, whereas lower-aged subjects taking GLP-1 RA (vs. DPP-4i) exhibited lower risk of “hospitalization for heart failure”. Moreover, older-aged subjects taking GLP-1 RA (vs. DPP-4i) had decreased risk of “acute myocardial infarction”, while middle-aged subjects taking GLP-1 RA (vs. DPP-4i) exhibited no evidence of a difference in treatment effect for risk of “acute myocardial infarction”. These findings align with current clinical guidelines, which recommend GLP-1 RA over DPP-4i for individuals at higher CV risk. While DPP-4i are generally considered to have neutral effects on cardiovascular outcomes, evidence from the SAVOR-TIMI 53 study suggests an increased risk of heart failure associated specifically with saxagliptin [10].

A consistent observation across the three age groups was that the relative reduction in hypoglycemia risk associated with initiation of DPP-4i, SGLT2i and GLP-1 RA compared to sulfonylureas became more pronounced with increasing age. These findings are in agreement with the current guidelines, which state that “older adults may be at higher risk of hypoglycemia” and as such sulfonylureas should be avoided, and in addition tight glycemic control may be less important than avoiding hypoglycemia [43,44].

We found significant differences across age groups for DPP-4i compared to sulfonylureas. For subjects on DPP-4i (vs. sulfonylureas), older-aged individuals showed no evidence of a difference in treatment effect for “acute renal failure”, whereas lower-aged individuals had a lower risk.

### 4.4 Sex Subgroup Heterogeneity

While prior research has largely overlooked sex-based heterogeneity, we found a significant difference in treatment effect between males and females. While females taking DPP-4i (vs. SGLT2i) showed improved “glycemic control”, males taking DPP-4i (vs. SGLT2i) exhibited poorer “glycemic control”. We found no publications comparing these two drug classes by sex; as such, the interpretation of this finding remains unclear.

### 4.5 Limitations

Despite the large number of T2D adults and robust data on many outcomes, the study has several limitations. First, although our study used rigorous methods to achieve balance between groups, we were unable to eliminate allocation bias, or systematic differences in baseline characteristics that can influence treatment assignment and outcomes [17]. Second, reliance on large administrative databases introduces complexity as the completeness and quality of data can vary across sources. Third, our analysis does not account for temporal variation in hazard ratios, which may lead to biased findings if summarized as a single average value [45]. Fourth, while several comparisons met the nominal significance threshold (p<0.05), we caution that such findings may reflect false positives due to the multiple hypothesis tests performed. Only one comparison met the conservative Bonferroni-corrected threshold (p<4.5×10^-5^), likely reflecting the limited statistical power to detect small subgroup effects. As such, these results should be interpreted as hypothesis-generating rather than conclusive evidence of subgroup-specific treatment effects. Fifth, while a multivariate model may have been able to detect higher-level heterogeneity of treatment effects across multiple factors (i.e. two-way interaction such as age and sex), these data were unavailable [46]. Finally, although our study examined heterogeneity of treatment effect across clinical and demographic subgroups, it did not address broader social determinants of health (e.g., race, ethnicity, income, educational level) or genetic, environmental and/or sociocultural factors that can influence drug metabolism, effectiveness and safety. To our knowledge, no large-scale comparative effectiveness data network captures comprehensive social determinants of health information; nonetheless, their absence remains an important consideration when interpreting the results.

### 4.6 Conclusions

In this evaluation of four glucose-lowering drug classes, we found one statistically significant difference and several hypotheses-generating nominally significant differences related to CV risk, renal impairment, age and sex. Overall, these findings can provide guidance to generate hypothesis-testing studies that will inform treatment decisions for T2D.

## CRediT author statement

**David M. Dávila-García:** Conceptualization, Methodology, Software, Validation, Formal analysis, Investigation, Resources, Data Curation, Writing – Original Draft, Writing – Review & Editing, Visualization, Project administration. **Thomas Falconer:** Methodology, Software, Formal analysis, Resources, Project administration. **Nicole Pratt:** Conceptualization, Methodology, Validation, Formal analysis, Writing – Review & Editing, Supervision, Project administration. **Karthik Natarajan:** Conceptualization, Validation, Resources, Writing – Review & Editing, Supervision, Project administration, Funding acquisition. **George Hripcsak:** Conceptualization, Methodology, Validation, Formal analysis, Investigation, Resources, Writing – Review & Editing, Visualization, Supervision, Project administration, Funding acquisition.

## Data Availability Statement

The data supporting the findings of this study are publicly available through the LEGEND-T2DM Evidence Explorer (https://data.ohdsi.org/LegendT2dmClassEvidenceExplorer/).

## Abbreviations

CKD: chronic kidney disease
CV: cardiovascular
DPP-4i: dipeptidyl peptidase-4 inhibitor
GLP-1 RA: glucagon-like peptide-1 receptor agonist
HR: hazard ratio
MACE: major adverse cardiovascular events
RI: renal impairment
SGLT2i: sodium-glucose cotransporter 2 inhibitor
T2D: type 2 diabetes
WRI: without renal impairment

---

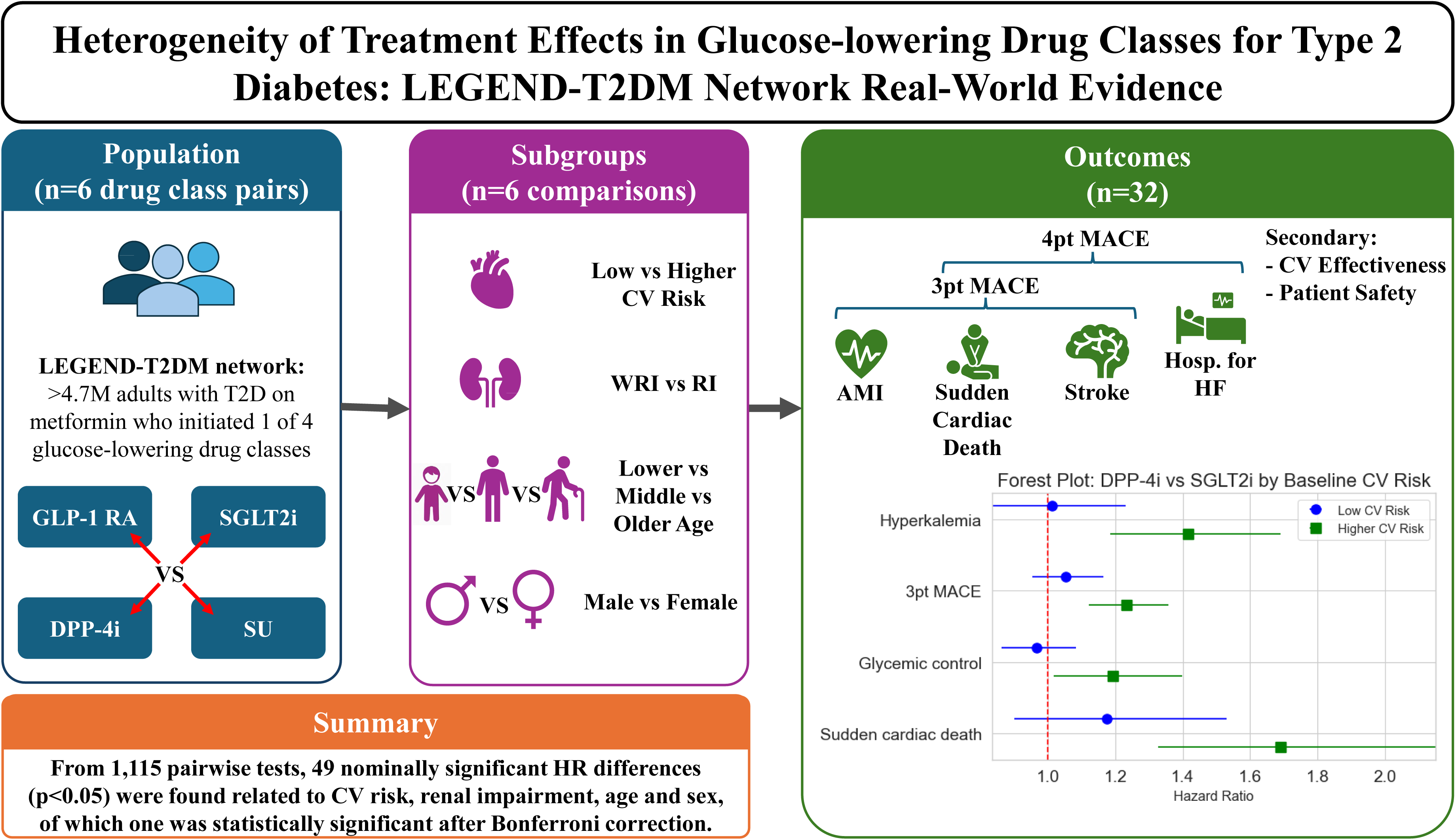

## REFERENCES

[1] Harding JL, Pavkov ME, Magliano DJ, Shaw JE, Gregg EW. Global trends in diabetes complications: a review of current evidence. Diabetologia 2019;62:3–16. 10.1007/s00125-018-4711-2.

[2] Global, regional, and national burden of diabetes from 1990 to 2021, with projections of prevalence to 2050: a systematic analysis for the Global Burden of Disease Study 2021. Lancet Lond Engl 2023;402:203–34. 10.1016/S0140-6736(23)01301-6.

[3] Rossing P, Caramori ML, Chan JCN, Heerspink HJL, Hurst C, Khunti K, et al. KDIGO 2022 Clinical Practice Guideline for Diabetes Management in Chronic Kidney Disease. Kidney Int 2022;102:S1–127. 10.1016/j.kint.2022.06.008.

[4] Abdul-Ghani M, DeFronzo RA, Del Prato S, Chilton R, Singh R, Ryder REJ. Cardiovascular Disease and Type 2 Diabetes: Has the Dawn of a New Era Arrived? Diabetes Care 2017;40:813–20. 10.2337/dc16-2736.

[5] Afkarian M, Sachs MC, Kestenbaum B, Hirsch IB, Tuttle KR, Himmelfarb J, et al. Kidney Disease and Increased Mortality Risk in Type 2 Diabetes. J Am Soc Nephrol JASN 2013;24:302–8. 10.1681/ASN.2012070718.

[6] Baumblatt J, Fryar C, Gu Q, Ashman J. Prevalence of Total, Diagnosed, and Undiagnosed Diabetes in Adults: United States, August 2021–August 2023. National Center for Health Statistics (U.S.); 2024. 10.15620/cdc/165794.

[7] Inoue K, Liu M, Aggarwal R, Marinacci LX, Wadhera RK. Prevalence and Control of Diabetes Among US Adults, 2013 to 2023. JAMA 2025. 10.1001/jama.2024.28513.

[8] Shi Q, Nong K, Vandvik PO, Guyatt GH, Schnell O, Rydén L, et al. Benefits and harms of drug treatment for type 2 diabetes: systematic review and network meta-analysis of randomised controlled trials. BMJ 2023;381:e074068. 10.1136/bmj-2022-074068.

[9] American Diabetes Association Professional Practice Committee. 9. Pharmacologic Approaches to Glycemic Treatment: Standards of Care in Diabetes—2025. Diabetes Care 2024;48:S181–206. 10.2337/dc25-S009.

[10] Scirica BM, Bhatt DL, Braunwald E, Steg PG, Davidson J, Hirshberg B, et al. Saxagliptin and Cardiovascular Outcomes in Patients with Type 2 Diabetes Mellitus. N Engl J Med 2013;369:1317–26. 10.1056/NEJMoa1307684.

[11] White WB, Cannon CP, Heller SR, Nissen SE, Bergenstal RM, Bakris GL, et al. Alogliptin after Acute Coronary Syndrome in Patients with Type 2 Diabetes. N Engl J Med 2013;369:1327–35. 10.1056/NEJMoa1305889.

[12] Green JB, Bethel MA, Armstrong PW, Buse JB, Engel SS, Garg J, et al. Effect of Sitagliptin on Cardiovascular Outcomes in Type 2 Diabetes. N Engl J Med 2015;373:232–42. 10.1056/NEJMoa1501352.

[13] Rosenstock J, Perkovic V, Johansen OE, Cooper ME, Kahn SE, Marx N, et al. Effect of Linagliptin vs Placebo on Major Cardiovascular Events in Adults With Type 2 Diabetes and High Cardiovascular and Renal Risk: The CARMELINA Randomized Clinical Trial. JAMA 2019;321:69–79. 10.1001/jama.2018.18269.

[14] Sahin I, Bakiner O, Demir T, Sari R, Atmaca A. Current Position of Gliclazide and Sulfonylureas in the Contemporary Treatment Paradigm for Type 2 Diabetes: A Scoping Review. Diabetes Ther 2024;15:1687–716. 10.1007/s13300-024-01612-8.

[15] Davies MJ, Aroda VR, Collins BS, Gabbay RA, Green J, Maruthur NM, et al. Management of hyperglycaemia in type 2 diabetes, 2022. A consensus report by the American Diabetes Association (ADA) and the European Association for the Study of Diabetes (EASD). Diabetologia 2022;65:1925–66. 10.1007/s00125-022-05787-2.

[16] Khera R, Aminorroaya A, Dhingra LS, Thangaraj PM, Pedroso Camargos A, Bu F, et al. Comparative Effectiveness of Second-Line Antihyperglycemic Agents for Cardiovascular Outcomes. J Am Coll Cardiol 2024;84:904–17. 10.1016/j.jacc.2024.05.069.

[17] Everett BM, Wexler DJ. Finding Truth in Observational and Interventional Studies in Diabetes and Cardiovascular Disease. J Am Coll Cardiol 2024;84:918–20. 10.1016/j.jacc.2024.06.028.

[18] Green JB, Everett BM, Ghosh A, Younes N, Krause-Steinrauf H, Barzilay J, et al. Cardiovascular Outcomes in GRADE (Glycemia Reduction Approaches in Type 2 Diabetes: A Comparative Effectiveness Study). Circulation 2024;149:993–1003. 10.1161/CIRCULATIONAHA.123.066604.

[19] Rosenstock J, Kahn SE, Johansen OE, Zinman B, Espeland MA, Woerle HJ, et al. Effect of Linagliptin vs Glimepiride on Major Adverse Cardiovascular Outcomes in Patients With Type 2 Diabetes: The CAROLINA Randomized Clinical Trial. JAMA 2019;322:1155–66. 10.1001/jama.2019.13772.

[20] Nair ATN, Wesolowska-Andersen A, Brorsson C, Rajendrakumar AL, Hapca S, Gan S, et al. Heterogeneity in phenotype, disease progression and drug response in type 2 diabetes. Nat Med 2022;28:982–8. 10.1038/s41591-022-01790-7.

[21] Khera R, Schuemie MJ, Lu Y, Ostropolets A, Chen R, Hripcsak G, et al. Large-scale evidence generation and evaluation across a network of databases for type 2 diabetes mellitus (LEGEND-T2DM): a protocol for a series of multinational, real-world comparative cardiovascular effectiveness and safety studies. BMJ Open 2022;12:e057977. 10.1136/bmjopen-2021-057977. [dataset]

[22] LEGEND-T2DM Evidence Explorer n.d. https://data.ohdsi.org/LegendT2dmClassEvidenceExplorer/ (accessed September 26, 2024).

[23] Suchard MA, Schuemie MJ, Krumholz HM, You SC, Chen R, Pratt N, et al. Comprehensive comparative effectiveness and safety of first-line antihypertensive drug classes: a systematic, multinational, large-scale analysis. The Lancet 2019;394:1816–26. 10.1016/S0140-6736(19)32317-7.

[24] Khera R, Dhingra LS, Aminorroaya A, Li K, Zhou JJ, Arshad F, et al. Multinational patterns of second line antihyperglycaemic drug initiation across cardiovascular risk groups: federated pharmacoepidemiological evaluation in LEGEND-T2DM. BMJ Med 2023;2:e000651. 10.1136/bmjmed-2023-000651.

[25] Schuemie MJ, Ryan PB, Pratt N, Chen R, You SC, Krumholz HM, et al. Principles of Large-scale Evidence Generation and Evaluation across a Network of Databases (LEGEND). J Am Med Inform Assoc JAMIA 2020;27:1331–7. 10.1093/jamia/ocaa103.

[26] Reich C, Ostropolets A, Ryan P, Rijnbeek P, Schuemie M, Davydov A, et al. OHDSI Standardized Vocabularies-a large-scale centralized reference ontology for international data harmonization. J Am Med Inform Assoc JAMIA 2024;31:583–90. 10.1093/jamia/ocad247.

[27] Hernán MA, Robins JM. Per-Protocol Analyses of Pragmatic Trials. N Engl J Med 2017;377:1391–8. 10.1056/NEJMsm1605385.

[28] Murray EJ, Caniglia EC, Swanson SA, Hernández-Díaz S, Hernán MA. Patients and investigators prefer measures of absolute risk in subgroups for pragmatic randomized trials. J Clin Epidemiol 2018;103:10–21. 10.1016/j.jclinepi.2018.06.009.

[29] Arnold SV, Inzucchi SE, Echouffo-Tcheugui JB, Tang F, Lam CSP, Sperling LS, et al. Understanding Contemporary Use of Thiazolidinediones: An Analysis From the Diabetes Collaborative Registry. Circ Heart Fail 2019;12:e005855. 10.1161/CIRCHEARTFAILURE.118.005855.

[30] Ryan PB, Buse JB, Schuemie MJ, DeFalco F, Yuan Z, Stang PE, et al. Comparative effectiveness of canagliflozin, SGLT2 inhibitors and non-SGLT2 inhibitors on the risk of hospitalization for heart failure and amputation in patients with type 2 diabetes mellitus: A real-world meta-analysis of 4 observational databases (OBSERVE-4D). Diabetes Obes Metab 2018;20:2585–97. 10.1111/dom.13424.

[31] Brody T. Chapter 9 - Biostatistics. In: Brody T, editor. Clin. Trials, Boston: Academic Press; 2012, p. 165–90. 10.1016/B978-0-12-391911-3.00009-8.

[32] Bewick V, Cheek L, Ball J. Statistics review 12: Survival analysis. Crit Care 2004;8:389–94. 10.1186/cc2955.

[33] Brønden A, Christensen MB, Glintborg D, Snorgaard O, Kofoed-Enevoldsen A, Madsen GK, et al. Effects of DPP-4 inhibitors, GLP-1 receptor agonists, SGLT-2 inhibitors and sulphonylureas on mortality, cardiovascular and renal outcomes in type 2 diabetes: A network meta-analyses-driven approach. Diabet Med J Br Diabet Assoc 2023;40:e15157. 10.1111/dme.15157.

[34] Xie Y, Bowe B, Xian H, Loux T, McGill JB, Al-Aly Z. Comparative effectiveness of SGLT2 inhibitors, GLP-1 receptor agonists, DPP-4 inhibitors, and sulfonylureas on risk of major adverse cardiovascular events: emulation of a randomised target trial using electronic health records. Lancet Diabetes Endocrinol 2023;11:644–56. 10.1016/S2213-8587(23)00171-7.

[35] American Diabetes Association Professional Practice Committee. 11. Chronic Kidney Disease and Risk Management: Standards of Care in Diabetes—2025. Diabetes Care 2024;48:S239–51. 10.2337/dc25-S011.

[36] Baksh SN, Segal JB, McAdams-DeMarco M, Kalyani RR, Alexander GC, Ehrhardt S. Dipeptidyl peptidase-4 inhibitors and cardiovascular events in patients with type 2 diabetes, without cardiovascular or renal disease. PloS One 2020;15:e0240141. 10.1371/journal.pone.0240141.

[37] Hanlon P, Butterly E, Wei L, Wightman H, Almazam SAM, Alsallumi K, et al. Age and Sex Differences in Efficacy of Treatments for Type 2 Diabetes: A Network Meta-Analysis. JAMA 2025. 10.1001/jama.2024.27402.

[38] American Diabetes Association Professional Practice Committee. 13. Older Adults: Standards of Care in Diabetes—2025. Diabetes Care 2024;48:S266–82. 10.2337/dc25-S013.

[39] Karagiannis T, Tsapas A, Athanasiadou E, Avgerinos I, Liakos A, Matthews DR, et al. GLP-1 receptor agonists and SGLT2 inhibitors for older people with type 2 diabetes: A systematic review and meta-analysis. Diabetes Res Clin Pract 2021;174. 10.1016/j.diabres.2021.108737.

[40] Zinman B, Wanner C, Lachin JM, Fitchett D, Bluhmki E, Hantel S, et al. Empagliflozin, Cardiovascular Outcomes, and Mortality in Type 2 Diabetes. N Engl J Med 2015;373:2117–28. 10.1056/NEJMoa1504720.

[41] Neal B, Perkovic V, Mahaffey KW, Zeeuw D de, Fulcher G, Erondu N, et al. Canagliflozin and Cardiovascular and Renal Events in Type 2 Diabetes. N Engl J Med 2017;377:644–57. 10.1056/NEJMoa1611925.

[42] Wiviott SD, Raz I, Bonaca MP, Mosenzon O, Kato ET, Cahn A, et al. Dapagliflozin and Cardiovascular Outcomes in Type 2 Diabetes. N Engl J Med 2019;380:347–57. 10.1056/NEJMoa1812389.

[43] McCoy RG, Swarna KS, Neumiller JJ, Polley EC, Deng Y, Mickelson MM, et al. Risk of Severe Hypoglycemia After Initiation of Noninsulin Glucose-Lowering Therapies in Adults With Type 2 Diabetes at Moderate Cardiovascular Disease Risk. Clin Diabetes 2024:cd240007. 10.2337/cd24-0007.

[44] Pilla SJ, Jalalzai R, Tang O, Schoenborn NL, Boyd CM, Golden SH, et al. A National Physician Survey of Deintensifying Diabetes Medications for Older Adults With Type 2 Diabetes. Diabetes Care 2023;46:1164–8. 10.2337/dc22-2146.

[45] Hernán MA. The Hazards of Hazard Ratios. Epidemiol Camb Mass 2010;21:13–5. 10.1097/EDE.0b013e3181c1ea43.

[46] Bellavia A, Murphy SA. Heterogeneity of Treatment Effects in Clinical Trials: Overview of Multivariable Approaches and Practical Recommendations. Circulation 2024;150:978–80. 10.1161/CIRCULATIONAHA.124.069857.

