## Supplementary Materials for "Heterogeneity of Treatment Effects of Glucose-lowering Drug Classes for Type 2 Diabetes: LEGEND-T2DM Network Real-World Evidence"

#### Study Design.

In this study, we leveraged LEGEND-T2DM (Large-scale Evidence Generation and Evaluation across a Network of Databases for Type 2 Diabetes Mellitus) network to obtain real-world data from over 4.7 million adults with T2D on metformin monotherapy who initiated one of four major glucose-lowering drug classes (glucagon-like peptide-1 receptor agonist [GLP-1 RA], sodium-glucose cotransporter 2 inhibitor [SGLT2i], dipeptidyl peptidase-4 inhibitor [DPP-4i] or sulfonylureas). The LEGEND-T2DM study implemented an active comparator, new-user cohort design to emulate a target trial comparing glucose-lowering drug classes [1–4]. Detailed study methodology are previously described in the LEGEND-T2DM study protocol [5].

The overall goal of the LEGEND framework is to systematically generate reliable real-world evidence, as further described by Schuemie et al. [6,7]. Specifically, the study applied a "federated analytical framework to address residual confounding, publication bias, and P hacking" [8–10]. This approach consisted of large-scale propensity score adjustment for measured confounding [11,12], a large set of negative control outcome experiments (n=100) to address unmeasured confounding and residual bias [13–16], cohort diagnostics to evaluate internal and external validity [3,17,18] and meta-analytic methods to aggregate evidence across data sources.

#### Propensity Score Adjustment.

Large-scale propensity score adjustment was performed to control for measured confounding and improve balance between cohorts for each pairwise drug class comparison and individual data sources [19]. Propensity scores were estimated using L1 regularized logistic regression with a broad range of predefined baseline patient characteristics (e.g. demographics, comorbidities, concomitant medication use, health care use prior to index date). Cox proportional hazards models were used to estimate hazard ratios of each outcome for each comparison, conditional on propensity score variable-ratio patient matching [20].

#### Negative Control Outcome Experiments.

Negative control outcome experiments were conducted to quantify and adjust for residual bias [21]. Negative control outcomes were selected based on the absence of known causal associations with the exposures under study (negative controls listed in Khera et al., Supplemental Table 4 [10]). For each drug class comparison and outcome, the null hypothesis of no treatment effect (hazard ratio = 1) was tested across the set of negative control outcomes. Deviations from the null were used to empirically estimate the distribution of systematic error (due to unmeasured confounding, measurement error and selection bias). The empirical null distribution was applied to calibrate the observed HR estimates and corresponding 95% confidence intervals [15].

#### Study Diagnostics.

While blinded to the results, study diagnostics were performed to evaluate the internal and external validity of findings for all comparisons. Diagnostics were conducted across three domains: a) evaluation of minimum detectable risk ratio (MDRR) as a metric for statistical power [22]; b) examination of preference score distributions between the target and comparator cohorts to assess empirical equipoise and generalizability; and c) assessment of covariate balance before and after propensity score adjustment, using absolute standardized mean differences (SMDs) [23].

Only data sources that met predefined diagnostic thresholds were included in the final analysis. Specifically, a study passed diagnostics if the MDRR was <4, >25% of subjects in each cohort had preference scores between 0.3 and 0.7, and the maximum SMD <0.15 after propensity score adjustment. All studies that passed diagnostics reported hazard ratio estimates, the corresponding 95% CIs, and p-values post-calibration.

#### Evidence Aggregation.

To synthesize evidence across non-overlapping data sources, calibrated hazard ratio estimates for each pairwise exposure comparison was aggregated using a random-effects meta-analysis. Evidence aggregation in this context aims to produce a summary effect estimate that accounts from both within- and between-data source variability. The random-effects model assumes that per-data source likelihoods are approximately normally distributed [24].

### References


[1] Yoshida K, Solomon DH, Kim SC. Active-comparator design and new-user design in observational studies. Nat Rev Rheumatol 2015;11:437–41. https://doi.org/10.1038/nrrheum.2015.30.

[2] Ryan PB, Schuemie MJ, Gruber S, Zorych I, Madigan D. Empirical Performance of a New User Cohort Method: Lessons for Developing a Risk Identification and Analysis System. Drug Saf 2013;36:59–72. https://doi.org/10.1007/s40264-013-0099-6.

[3] Schuemie MJ, Cepeda MS, Suchard MA, Yang J, Tian Y, Schuler A, et al. How Confident Are We About Observational Findings in Health Care: A Benchmark Study. Harv Data Sci Rev 2020;2. https://doi.org/10.1162/99608f92.147cc28e.

[4] Concato J, Shah N, Horwitz RI. RANDOMIZED, CONTROLLED TRIALS, OBSERVATIONAL STUDIES, AND THE HIERARCHY OF RESEARCH DESIGNS. N Engl J Med 2000;342:1887–92.

[5] Khera R, Schuemie MJ, Lu Y, Ostropolets A, Chen R, Hripcsak G, et al. Large-scale evidence generation and evaluation across a network of databases for type 2 diabetes mellitus (LEGEND-T2DM): a protocol for a series of multinational, real-world comparative cardiovascular effectiveness and safety studies. BMJ Open 2022;12:e057977. https://doi.org/10.1136/bmjopen-2021-057977.

[6] Schuemie MJ, Ryan PB, Pratt N, Chen R, You SC, Krumholz HM, et al. Principles of Large-scale Evidence Generation and Evaluation across a Network of Databases (LEGEND). J Am Med Inform Assoc 2020;27:1331–7. https://doi.org/10.1093/jamia/ocaa103.

[7] Schuemie MJ, Ryan PB, Pratt N, Chen R, You SC, Krumholz HM, et al. Large-scale evidence generation and evaluation across a network of databases (LEGEND): assessing validity using hypertension as a case study. J Am Med Inform Assoc JAMIA 2020;27:1268–77. https://doi.org/10.1093/jamia/ocaa124.

[8] Frieden TR. Evidence for Health Decision Making — Beyond Randomized, Controlled Trials. N Engl J Med 2017;377:465–75. https://doi.org/10.1056/NEJMra1614394.

[9] Sherman RE, Anderson SA, Pan GJD, Gray GW, Gross T, Hunter NL, et al. Real-World Evidence — What Is It and What Can It Tell Us? N Engl J Med 2016;375:2293–7. https://doi.org/10.1056/NEJMsb1609216.

[10] Khera R, Aminorroaya A, Dhingra LS, Thangaraj PM, Pedroso Camargos A, Bu F, et al. Comparative Effectiveness of Second-Line Antihyperglycemic Agents for Cardiovascular Outcomes. J Am Coll Cardiol 2024;84:904–17. https://doi.org/10.1016/j.jacc.2024.05.069.

[11] Tian Y, Schuemie MJ, Suchard MA. Evaluating large-scale propensity score performance through real-world and synthetic data experiments. Int J Epidemiol 2018;47:2005–14. https://doi.org/10.1093/ije/dyy120.

[12] Rosenbaum PR, Rubin DB. The central role of the propensity score in observational studies for causal effects n.d.

[13] Schuemie MJ, Ryan PB, DuMouchel W, Suchard MA, Madigan D. Interpreting observational studies: why empirical calibration is needed to correct p-values. Stat Med 2014;33:209–18. https://doi.org/10.1002/sim.5925.

[14] Schuemie MJ, Hripcsak G, Ryan PB, Madigan D, Suchard MA. Robust empirical calibration of p-values using observational data. Stat Med 2016;35:3883–8. https://doi.org/10.1002/sim.6977.

[15] Schuemie MJ, Hripcsak G, Ryan PB, Madigan D, Suchard MA. Empirical confidence interval calibration for population-level effect estimation studies in observational healthcare data. Proc Natl Acad Sci U S A 2018;115:2571–7. https://doi.org/10.1073/pnas.1708282114.

[16] Hwang H, Quiroz JC, Gallego B. Assessing the effectiveness of empirical calibration under different bias scenarios. BMC Med Res Methodol 2022;22:208. https://doi.org/10.1186/s12874-022-01687-6.

[17] Schuemie MJ, Ryan PB, Hripcsak G, Madigan D, Suchard MA. Improving reproducibility by using high-throughput observational studies with empirical calibration. Philos Trans R Soc Math Phys Eng Sci 2018;376:20170356. https://doi.org/10.1098/rsta.2017.0356.

[18] Conover MM, Ryan PB, Chen Y, Suchard MA, Hripcsak G, Schuemie MJ. Objective study validity diagnostics: a framework requiring pre-specified, empirical verification to increase trust in the reliability of real-world evidence. J Am Med Inform Assoc JAMIA 2025;32:518–25. https://doi.org/10.1093/jamia/ocae317.

[19] Zhang L, Wang Y, Schuemie MJ, Blei DM, Hripcsak G. Adjusting for Indirectly Measured Confounding Using Large-Scale Propensity Score. J Biomed Inform 2022;134:104204. https://doi.org/10.1016/j.jbi.2022.104204.

[20] Austin PC. The use of propensity score methods with survival or time-to-event outcomes: reporting measures of effect similar to those used in randomized experiments. Stat Med 2014;33:1242–58. https://doi.org/10.1002/sim.5984.

[21] Lipsitch M, Tchetgen ET, Cohen T. Negative Controls: A Tool for Detecting Confounding and Bias in Observational Studies. Epidemiol Camb Mass 2010;21:383–8. https://doi.org/10.1097/EDE.0b013e3181d61eeb.

[22] Suchard MA, Schuemie MJ, Krumholz HM, You SC, Chen R, Pratt N, et al. Comprehensive comparative effectiveness and safety of first-line antihypertensive drug classes: a systematic, multinational, large-scale analysis. The Lancet 2019;394:1816–26. https://doi.org/10.1016/S0140-6736(19)32317-7.

[23] Austin PC. Balance diagnostics for comparing the distribution of baseline covariates between treatment groups in propensity-score matched samples. Stat Med 2009;28:3083–107. https://doi.org/10.1002/sim.3697.
[24] DerSimonian R, Laird N. Meta-analysis in clinical trials. Control Clin Trials 1986;7:177–88. https://doi.org/10.1016/0197-2456(86)90046-2.
